# Implementation of the Liberia National Community Health Assistant (NCHA) Program and Under-five Mortality: A study protocol

**DOI:** 10.1101/2022.07.15.22277669

**Authors:** Dominik Jockers, Richard Ngafuan, Till Baernighausen, Albert Kessley, Emily E. White, Avi Kenny, John Kraemer, John Geedeh, Jeffrey Rozelle, Leah Holmes, Hawa Obaje, Sylvester Wheh, Jon Pederson, Mark J. Siedner, Savior Mendin, Marion Subah, Lisa R Hirschhorn

## Abstract

**Introduction:** Between 2018 and 2022 the Liberian Government implemented the National Community Health Assistant (NCHA) program to improve provision of maternal and child health care to underserved rural areas of the country. Whereas the contributions of this and similar community health worker (CHW) based healthcare programs have been associated with improved process measures, the impact of a governmental CHW program at scale on child mortality has not been fully established.

**Methods/Design:** We will conduct a cluster sampled, community-based survey with landmark event calendars to retrospectively assess child births and deaths among all children born to women in the Grand Bassa District of Liberia. We will use a mixed effects discrete survival model, taking advantage of the staggered program implementation in Grand Bassa districts over a period of 4 years to compare rates of under-5 child mortality between the pre- and post-NCHA program implementation periods.

**Discussion:** This study will be the first to estimate the impact of the Liberian NCHA program on under-5 mortality.

**Trial registration:** clinicaltrials.gov NCT: NCT05123378

## Introduction

Under-5 mortality (U5M) has been reduced globally from 93 deaths per 1,000 live births in 1990 to 38 per 1,000 live births in 2019. Nevertheless, 1 in 13 children continue to die before their fifth birthday in sub-Saharan Africa. According to the World Health Organization, most of the deaths are due to preventable and treatable diseases (WHO, 2017).

Liberia ranks among the worst nations globally in child health outcomes, with U5M rate estimated at 93 per 1000 live births (Liberia Institute of Statistics and Geo-Information Services - LISGIS et al., 2021). Remote areas are facing the greatest burden of mortality because of poor access and utilization of health care services (Kentoffio et al., 2016). The primary causes of U5M in the region are malaria, diarrhea and acute respiratory infections (ARI) (Liu et al., 2015; Simen-Kapeu et al., 2021), which are all preventable and treatable with health interventions that are feasible for implementation in low-income countries (Jones et al., 2003).

Many countries have adopted integrated Community Case Management (iCCM) strategies to address the causes and reduce U5M. Community health workers (CHWs) are at the core of many iCCM strategies. They act within their local community and provide services such as treatment of uncomplicated childhood diseases and referring complicated cases to health facilities (Perry et al., 2014). iCCM interventions have been shown to reduce U5M due to an increased treatment of malaria, ARI and diarrhea (Baqui et al., 2002; Das et al., 2013; Delacollette et al., 1996; Nyqvist et al., 2019; Sazawal & Black, 2003; Theodoratou et al., 2010; Victora et al., 2000).

In June, 2016, the Liberian Ministry of Health (MOH) initiated a national professionalized, incentivized, and clinically supervised community health worker cadre known as Community Health Assistants (CHA). These CHAs are embedded within the public-sector health workforce, with the goal of extending the reach of the country’s primary health care system to an estimated 1.2 million people through a standardized national community health model. The CHAs provide a package of health care services and epidemic surveillance within communities and households on an equitable basis.

Although numerous studies have demonstrated improvements in child outcomes with iCCM interventions, evidence around the effects of government-run, CHW-delivered healthcare on child mortality in rural areas of sub-Saharan Africa is limited. Our central hypothesis is that the National Community Health Assistant (NCHA) program in Liberia reduced U5M in rural Liberia. To test this hypothesis, we will conduct a large, population-representative survey across a major county in Liberia, and use a mixed effects discrete survival model to retrospectively compare mortality in the pre- and post-implementation periods in order to estimate the effect of the CHA program on U5M.

## Method

### Study setting and population

Data collection did start in Grand Bassa county in January 2022. The county encompasses an area of 7,936 square kilometers and had an estimated total population of 224,839 in 2008 (LISGIS, 2009). The target population of the NCHA are households in communities with a distance of more than 5 km from the nearest health facility. Based on a county-wide community GIS mapping exercise conducted by LMH immediately before program implementation, 1,733 communities were identified as remote, totaling 23,702 households and approximately 13,400 people and 20,900 children under the age of five years. The remote population is predominantly poor and levels of education are low. In Table A1 in the Appendix we show descriptive statistics for the study population. The study has been registered at clinicaltrials.gov [NCT: NCT05123378]. Approval for the study was obtained from the University of Liberia-Pacific Institute for Research and Evaluation Institutional Review Board (18^th^ of November 2021, 18-11-140) and Advarra Institutional Review Board (25^th^ of January 2021, Pro00048901), United States.

### Community Health Worker Intervention

#### The Program

As part of the post-Ebola investment plan for building a resilient health system in Liberia 2015-2021, the MOH prioritized the establishment of a robust cadre of community health workers in remote communities that were situated more than 5 km from the nearest health facility. Through the NCHA program, members of remote communities select their CHA. Those selected by their communities have to fulfil eligibility requirements, including being permanent residents in the community, aged 18 to 50 years, possess basic writing and math skills, be physically and mentally fit and must be fluent in the dialect spoken in their community. Once selected, CHAs complete a standardized training curriculum in community health and surveillance, child health and maternal and neonatal health. Services are covering treatment of diarrhea, ARI, and malaria following iCCM protocols, as well as family planning and birth preparedness. Since 2020 COVID-education and -referral has also been included in the CHAs work. Everyone in the communities more than 5 km from the nearest health facility is eligible to use the program directly or indirectly. CHAs are compensated with a monthly cash incentive of US $70 for about 20 hours of work per week. Program implementation is preceded by community engagement activities, which involves obtaining the support and advice of community leaders.

#### Implementation

The Liberian NCHA program was initiated in a stepwise manner across the eight districts of Grand Bassa County by the government of Liberia beginning in March 2018. It is planned to conclude in June 2022 (Figure 1). We will take advantage of this staggered implementation to form a study design similar in structure to a stepped wedge cluster randomized trial. However, the order of implementation was not randomized; judged by the Liberian MOH districts with more need for health interventions were prioritized in the intervention roll out. The staggering of implementation was conducted for operational and logistical reasons, rather than to facilitate an evaluation.

**Figure 1:**
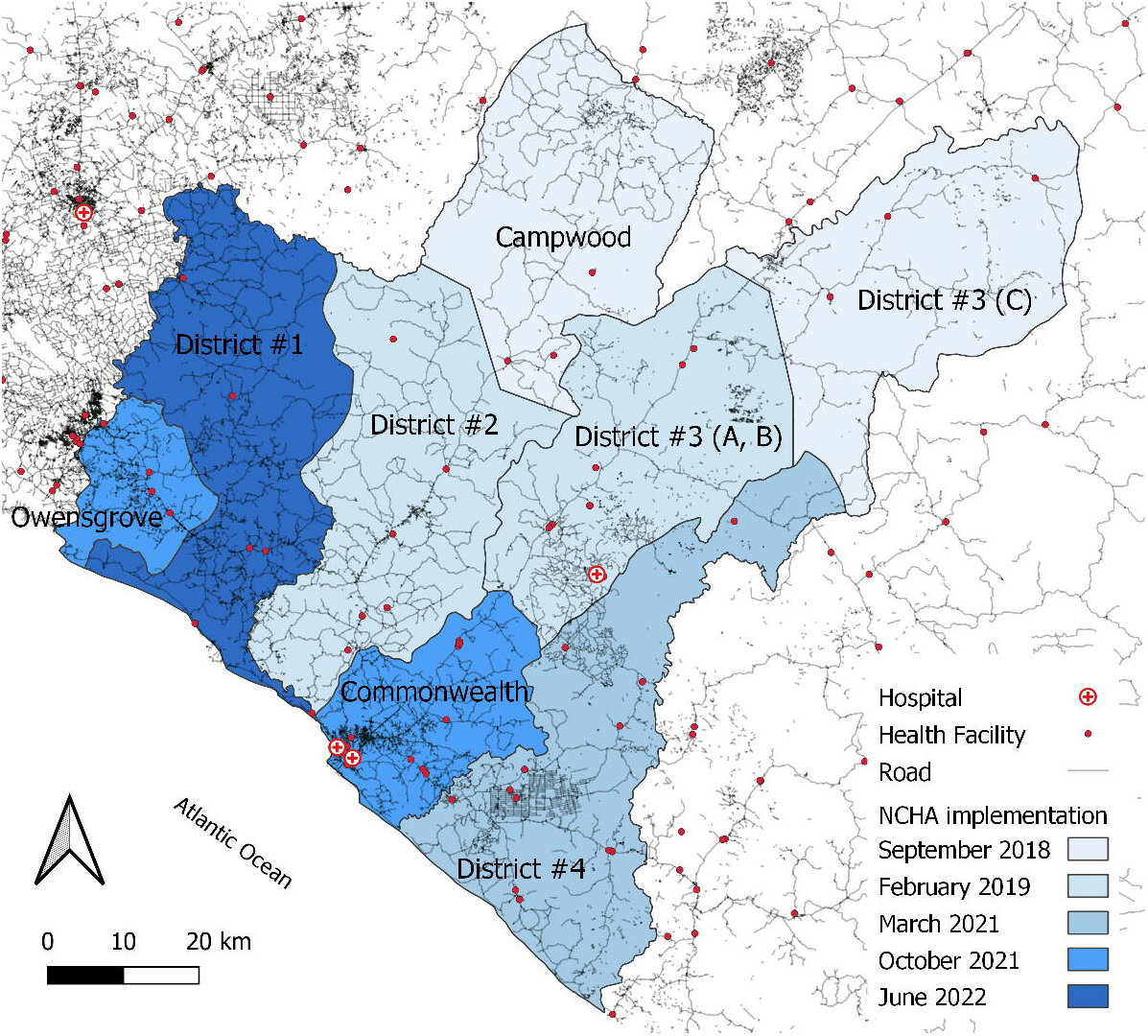
National Community Health Assistant (NCHA) program implementation schedule in Grand Bassa, Liberia Source: Dominik Jockers

### Study design

Starting in January 2022, we will conduct a census to assess U5M among the children of all women who complete the survey. The study is registered at clinicaltrials.gov (NCT05123378).

### Data collection

#### Impact Assessment Survey

In the 2022 impact assessment survey, we will collect data on births and child deaths from mothers aged 15 to 54 in all eight health districts of Grand Bassa (see Appendix Table A2). We will use a survey tool based on the validated Demographic and Health Survey (DHS) that includes a full birth history, as well as questions on socioeconomic status, education, and covariates that we expect to be associated with our endpoints. We will supplement this dataset with geographic and community-level covariates obtained through a separate district mapping process. Whenever possible, the survey will be conducted in Liberian English. If a respondent speaks only Bassa, a local dialect without a commonly used written form, the enumerator will do on-the-spot translation, which they will be trained to do. We will obtain death dates using the standard Demographic and Health Survey approach, augmented with recall-enhanced elicitation. For the recall-enhanced elicitation, mothers are asked to recall all their children death dates if occurred within the last four years. The same method has been applied in similar settings by Nyqvist et al. (2019). Anchoring methods with landmark events will be used in the questionnaire to foster the outcome documentation (Loftus & Marburger, 1983). We will use a calendar developed specific to Grand Bassa County, providing landmark events helping to clearly mark the reference period. Reluctance to report U5M is not expected to vary systematically by program implementation date.

### Survey sample selection

The impact assessment survey will be a full census; all 1,733 remote target communities in Grand Bassa will be selected and all households within selected communities will be surveyed. Within each household the survey population will consist of all women of reproductive age who are at least 15 years old (15-54). We selected the sample size to power our analysis using a mixed effects discrete survival model based on the assumption of a 25% decrease in the hazard of U5M following program implementation (see power calculation below).

### Statistical Analysis

The Liberian NCHA intervention and data collection will provide a survival data set for U5M at the community level, covering a time period from January 2018 till August 2022. To estimate program effects on U5M we will use a mixed effects discrete survival model.

Our primary outcome of interest will be U5M and our primary exposure of interest will be period will be presence of the NCHA program. Our model will account for the possibility of a time-varying treatment effect. Children will be included in the risk set from their date of birth or January 2018 (starting date of the panel), whichever is later, until their death, fifth birthday or date of the survey in 2022. Children born before implementation who remain under observation at the implementation date in their community will provide observation time to both the pre- and post-observation time periods. We will fit a mixed effects survival model with a random effect at the community level to estimate the effect of the NCHA program on mortality. To mitigate bias due to unobserved time-invariant confounders that were caused by the non-randomized implementation schedule of the program, we will include a fixed effect at the district level. We will also control for baseline demographic and geographic covariates.

### Power Calculation

We chose to perform a full census of the district’s 1,733 remote communities based on a set of simulation-based power calculations. Briefly, this involved simulating the population of eligible women (including their complete birth histories), taking a complex sample from this population, and running the planned statistical analysis on this sample. This entire procedure was performed repeatedly, and power was estimated as the percentage of simulation replicates for which the null hypothesis was rejected. Power was estimated as 82% for a full census. The treatment effect was assumed to be a 25% decrease in the hazard of U5M, linearly decreasing from the start of the program to six months and constant thereafter. The baseline mortality rate was set at roughly 100 deaths per 1,000 live births. The simulated dataset included random intercepts at the level of the community and at the level of the community-treatment interaction, allowing for heterogeneity in terms of both the baseline mortality rate and the effectiveness of the intervention.

## Discussion

This study will estimate the impact of Liberian NCHA program including iCCM and perinatal care support in remote Liberia on U5M. The staggered implementation of scale up within Grand Bassa offers a unique opportunity to evaluate the effectiveness of this program on a population scale. The analysis will allow us to evaluate the implementation of a critical healthcare delivery program by the Liberian Ministry of Health on U5M within the target communities.

There are multiple unique aspects of this study which will enhance both its scope and generalizability. The total population census with a program coverage of 1.753 communities and 23,702 households in the Grand Bassa county of Liberia, we will take advantage of a notably large sample size. The fact that the NCHA program is being implemented staggered across eight study health districts in four phases also gives us the opportunity to apply mixed effects discrete survival model with a random effect by community and district fixed effects to compare the intervention effect both within and between geographic regions. Finally, the use of U5M as the primary outcome, as opposed to intermediate health care delivery indicators and process measures, will allow us to measure the population level impacts of such a CHW-based child health care program at scale.

There are limitations to the design of this study. First, the order of implementation by district was non-randomized. Because the districts with the worst health outcomes were targeted by the intervention first, our estimate of the intervention effect estimate may be biased. Nonetheless, the stepped approach to implementation, availability of pre- and post-implementation data in the districts, and our use of district fixed effects and community covariates all enhance the causal inference of this study design. Second, the random effects at the community level rely on an assumption of minimal correlation of unobserved confounders with NCHA program exposure. Third, the data for our outcome relies on endline survey data, thus selective migration by intervention implementation date potentially causes bias in the main estimates. To address this, we will include a sensitivity analysis that limits the analysis only to those mothers who report living in the implementation area for the full study period. Third, U5M is assessed in the survey design by recall of such events in the last four years. There may be bias associated with mothers not remembering the correct birth/death dates of children. We will assume that this recall bias (and related biases, e.g. mothers’ reluctance to discuss child deaths) is minimal and does not differ significantly by geography or intervention status date. To mitigate to this source of bias and increase the validity of death reporting, we will employ a landmark event calendar, which has been previously shown to improve precision of event reporting (Loftus & Marburger, 1983). Finally, the fidelity of implementation is not a component of this impact study, but is the focus of ongoing program evaluation to measure dose, coverage and effect of contextual factors including health facility readiness and the COVID-19 pandemic.

Prior studies have demonstrated improvement in process measures from implementation of small scale governmental CHW programs in similar settings including Liberia (Fiori et al., 2021; White et al., 2018). Whereas Nyqvist et al. (2019) were able to randomize program exposure across communities, the program was different from our evaluation in that it is was operated by a non-governmental organization. Our study will add to the literature by providing evidence from a large scale, governmental-led CHW program.

In summary, this study will provide invaluable insight on the impact of large-scale, publicly operated CHW programs on child health. Results will be directly applicable both to the Liberian Ministry of Health, as well as to other governmental entities supporting similar CHW-based rural child health programs in the region. Because such rural areas are known to face the largest U5M burden in many sub-Saharan countries, our study results will directly respond to the need to understand the optimal methods of improving health care among many of the most vulnerable populations in the region.

## Supporting information

SPIRIT checklist

## Data Availability

The datasets used and analyzed during the current study are available from the corresponding author on reasonable request.

## Abbreviations

ARI: acute respiratory infections
iCCM: integrated Community Case Management
CHA: Community Health Assistants
CHW: community health worker
DHS: Demographic and Health Survey
MOH: Ministry of Health
NCHA: National Community Health Assistant
U5M: Under-5 mortality

## Declarations

### Ethics approval and consent to participate

Approval for the surveys was obtained from the University of Liberia-Pacific Institute for Research and Evaluation Institutional Review Board (18-11-140) and Advarra Institutional Review Board (Pro00048901), United States. All participants gave [verbal/written] informed consent to participate. Written informed consent was obtained from a parent or guardian for participants under 16 years old.

### Consent for publication

Not applicable

### Authors’ contributions

LH is Principal Investigator (PI) and responsible for overall conceptualization of study design. MS is Partner PIs and responsible for design and implementation in Grand Bassa, Liberia. DJ led the drafting of the manuscript. AK, JK, and MJS contributed to conceptualization and study design. AK, JK, MJS, TB and DJ were responsible for statistical method development. AK provided the power calculation. EW, AK, JK, LH, HO, JP and MS provided technical input. RN, AK, JG, SW and SM contributed with local context expertise; HO coordinated between contributors. All authors reviewed a draft and read and approved the final manuscript.

## Acknowledgements

Not Applicable

## Supporting Information

Document 1: Appendix

Document 2: SPIRIT checklist

